# Diagnostic accuracy of the Point-of-Care Standard G6PD test™ (SD Biosensor) for Glucose-6-phosphate dehydrogenase deficiency: a systematic review of the literature

**DOI:** 10.1101/2024.02.10.24302326

**Authors:** Juan Camilo Martínez, Viviana Vélez-Marín, Mary Lopez-Perez, Daniel Felipe Patiño, Ivan D. Florez

## Abstract

Glucose-6-Phosphate Dehydrogenase deficiency (G6PDd) is a common genetic enzymopathy that can induce hemolysis triggered by various factors, including some anti-malarial drugs. Although many Point-of-Care (PoC) tests, such as STANDARD G6PD^TM^ manufactured by SD biosensor (StandG6PD-BS), are available to detect G6PDd, its pooled diagnostic test accuracy (DTA) remains unknown. To estimate the DTA of StandG6PD-BS at various thresholds of G6PDd, we conducted a systematic review with a DTA meta-analysis, searching EMBASE, MEDLINE, and SciELO databases up to June 30, 2023. We included studies measuring G6PD activity using StandG6PD-BS (reference test) and spectrophotometry (gold standard) in patients suspected of having G6PDd. We assessed the risk of bias (RoB) of the studies using the QUADAS-2 tool and the certainty of evidence (CoE) with the GRADE approach. Our approach included the estimation of within-study DTA, a random-effect bivariate meta-analysis to determine the pooled sensitivity and specificity for 30%, 70%, and 80% enzyme levels’ thresholds, and a graphical analysis of the heterogeneity using crosshair and Confidence Regions on receiver operating characteristic (ROC) space plots. After screening 2,407 reports, we included four studies with 7,864 participants covering all thresholds. Two studies had high RoB in QUADAS-2 domains 2 and 3, and the others had low RoB. We also found low, moderate, and high heterogeneity at the 30%, 70%, and 80% thresholds, respectively. The pooled sensitivity was 99.1% (95%CI 96.9-99.7%, CoE: high), 95.7% (92.0-97.0%, high), and 90% (78.0-96.5%, low) for 30%, 70%, and 80% thresholds, respectively. The pooled specificity was 97.4% (95%CI 95.0; 98.4%, high); 92.9% (85.0-96.4%, high); and 89.0% (76.0-96.0%, moderate) for 30%, 70%, and 80% thresholds, respectively. In conclusion, StandG6PD-BS is a PoC test with high sensitivity and specificity to detect G6PDd at different thresholds.

**Author summary:** Glucose-6-Phosphate Dehydrogenase deficiency (G6PDd) is a common genetic disease that can induce the destruction of red blood cells leading to anemia triggered by various factors, including some drugs used for malaria treatment. After a literature search in different databases up to January 31, 2023, we pooled diagnostic test accuracy of the Point-of-Care (PoC) STANDARD G6PDTM test manufactured by SD biosensor (StandG6PD-BS) used to identify the G6PDd. Although two of the four studies included showed a high Risk of Bias related to the index test and the reference standard domains of the QUADAS-2 tool, the pooled sensitivity and specificity for 30%, 70%, and 80% enzyme levels’ thresholds were around 90%, with better sensitivity and specificity values for the 30% threshold (99.1% and 97.4%) compared with 70% (95.7% and 92.9%) and 80% (90% and 89%) threshold. We found low, moderate, and high heterogeneity at the 30%, 70%, and 80% thresholds. In conclusion, StandG6PD-BS is a PoC test with high sensitivity and specificity to detect G6PDd at different thresholds.

## Introduction

Glucose-6-phosphate dehydrogenase deficiency (G6PDd) is a genetic disorder linked to the X chromosome, with hemizygous males and homozygous females having a deficient activity phenotype (< 30% of enzyme levels). In contrast, heterozygous females may have a normal (> 80%) or intermediate activity phenotype (30% to 80% enzyme level) (1). Individuals with deficient activity may experience hemolytic episodes triggered by intrinsic or extrinsic factors such as medications or certain foods (2). G6PDd is the most common enzymopathy in humans, with variable frequencies and distinctive region-specific distribution (3). It is particularly common in malaria-endemic regions (1,4), with an estimated frequency of 8%-10% (∼350 to 400 million cases per year) and over 200 identified genetic polymorphisms (5). This overlapping is attributed to the protective effect of G6PDd against malaria (1).

Malaria caused by *Plasmodium vivax* is geographically widespread and counts for most cases outside Africa, particularly in the Americas and South-East Asia (6). Primaquine (PQ) and the novel tafenoquine (TQ) are the only two approved drugs for treating hepatic stages and are used for the radical cure of uncomplicated malaria by *P. vivax* (5,7). However, these drugs may precipitate hemolytic crises in individuals with G6PDd (1,5,7), with the severity of the reaction being proportional to the dose of the medication received and the enzyme genotype (5,7). To ensure safe administration of these drugs, the World Health Organization (WHO) recommends testing for G6PDd in those requiring treatment with PQ (7). This could be achieved by using Point-of-Care (PoC) tests to determine G6PD activity before administering PQ and TQ, as has been recommended by multiple studies (8).

Several tests are currently available to determine the G6PD activity, but they have different performance (9,10). Since the gold standard for G6PD measurement (spectrophotometry) is not suitable for PoC testing, qualitative tests have been developed with variable diagnostic performance and operational characteristics (11–13). While those tests discriminate between normal and deficient G6PD activity and therefore are sufficient to guide PQ treatment, they may not be dependable enough to prevent drug-induced hemolysis with the introduction of TQ. Consequently, more reliable diagnostic tests are required, and one of such test is the semi-quantitative assay STANDARD G6PD^TM^ (SD biosensor, Republic of Korea), an enzymatic colorimetric assay intended to aid the detection of G6PDd (14). This PoC test provides a numeric measurement of G6PD enzymatic activity and total hemoglobin (Hb) concentration in fresh capillary and venous human whole blood specimens (14) and allows classification of the G6PD activity as deficient, intermediate, or normal according to thresholds provided by the manufacturer (14).

Despite the recommendation for PoC quantitative or semi-quantitative testing before administering antimalarial treatment, there is only one synthesis of the diagnostic test accuracy (DTA) of the STANDARD G6PD^TM^ test manufactured by SD Biosensor (StandG6PD-BS) (15) in which the authors pooled the individual results of various studies without a complete systematic review approach, risk of bias, and certainty of evidence assessment. In this systematic review and metanalysis, we estimated the pooled DTA of the StandG6PD-BS for different thresholds of G6PD activity.

## Materials and methods

### Protocol and registration

This systematic review and DTA meta-analysis were conducted and reported following the Preferred Reporting Items for Systematic Reviews and Meta-analyses of DTA Studies (PRISMA-DTA) guidelines. The protocol was registered in the PROSPERO database (CRD42022311085).

### Search and study selection

Two authors (JCM, VVM) performed a structured search in MEDLINE (via PubMed), EMBASE, and SciELO databases on August 2^nd^, 2022, and updated it on January 31, 2023, and on June 30, 2023 without language or date restrictions. Our search strategy is outlined in **Table S1**. We included prospective or retrospective studies that measured G6PD activity levels using the reference standard enzymatic test (spectrophotometry G6PD assay) and the reference test (StandG6PD-BS). The studies must have reported enough data to calculate diagnostic performance measures, i.e., true positives (TP), true negatives (TN), false positives (FP), and false negatives (FN), for at least one threshold (30%, 70%, and 80%) of G6PD activity.

Using Rayyan software (Rayyan QCRI, Qatar) (16), two authors screened titles and abstracts retrieved from searches, and only those records considered eligible by both reviewers were retrieved in full texts for the next stage. The same authors (JCM, VVM) reviewed the potentially eligible full texts, independently and in duplicate, based on the pre-specified inclusion criteria. Those studies considered eligible by both reviewers were included. Disagreements were resolved by consensus among the reviewers and with the participation of a third reviewer if needed (MLP, IDF).

The same authors (JCM, VVM) independently and in duplicate extracted the data from the included studies using a prespecified data extraction form designed in Google Forms (Google LLC, US), which was discussed and piloted among the research team. For each study, we extracted the following information: first author, year of publication, title, population, data for every threshold recommended by the manufacturer (SD biosensor®), number of participants, age (mean and standard deviation), sex, type of blood sample (venous or capillary), and the data needed for 2×2 contingency tables (TP, FP, FN, and TN). Disagreements in the data extraction process were discussed between the reviewers.

### Risk of bias assessment and certainty of the evidence

Two authors (JCM, VVM) independently assessed the Risk of Bias (RoB) for the included studies using the Quality Assessment of Diagnostic Accuracy Studies 2 (QUADAS-2) tool (17). The QUADAS-2 instrument evaluates the RoB of DTA studies with four domains (patient selection, index test, reference standard, and flow and timing), judging each as high, low, or unclear in risk of bias and concerns regarding applicability. The authors discussed any disagreements and resolved them through consensus.

We assessed the Certainty of Evidence (CoE) using the GRADE framework for DTA systematic reviews (18,19). This approach evaluates four criteria: RoB (judged using the QUADAS-2 assessment), indirectness, inconsistency, publication bias and impression, and rates the certainty of the evidence in high, moderate, low, and very low.

### Statistical analysis

We used R Software and the package *mada* (version 4.1.2, The R Foundation for Statistical Computing) for the statistical analyses (20) and the GRADE Pro GDT platform (McMaster University 2015, developed by EvidencePrime, Inc) for creating the Summary of Findings (SoF) tables. First, we calculated sensitivity, specificity, positive likelihood ratio (+LR), negative likelihood ratio (-LR), and diagnostic odds ratio (DOR) at every threshold. The cut-off recommended for the StandG6PD-BS test manufacturer is a valuable tool for therapeutic decisions because enzyme levels define the use of specific treatment. Thus, enzymatic levels above 30% allow using PQ and other drugs at specific doses; levels above 70% allow using TQ and any PQ treatment schedule; and levels above 80% define a normal G6PD activity. Then, we fitted the type of blood sample data at each threshold with the *Reistma* model (21), a bivariate random-effect meta-analysis. With this approach, we calculated the pooled sensitivity, specificity, +LR, and –LR and their corresponding 95% confidence intervals (95% CI) utilizing model estimations. We used data only from female participants at 70% and 80% thresholds. We pooled the results for venous blood samples because the combined capillary samples were too small and not measured in all the studies.

For between-studies heterogeneity, we used visual inspection of forest plots for DTA pooled measures, crosshair plots (sensitivity vs. false positive rate), confidence regions in the *Receiver Operating Characteristic (*ROC) space, evaluating the overlap in the confidence intervals, and *chi-square* test for homogeneity (*p-*values <0.05 were considered significant, and therefore as with heterogeneity). We did not obtain enough studies to fit the sensitivity analysis models as stated in the registered protocol.

## Results

### Search results

We obtained 2,818 records and after removing duplicates, we screened 2,407 unique reports and identified nine potentially eligible studies. After the full-text assessment, we excluded five and included four studies that met the inclusion criteria for qualitative and quantitative synthesis (**Figure 1**).

**Figure 1.**
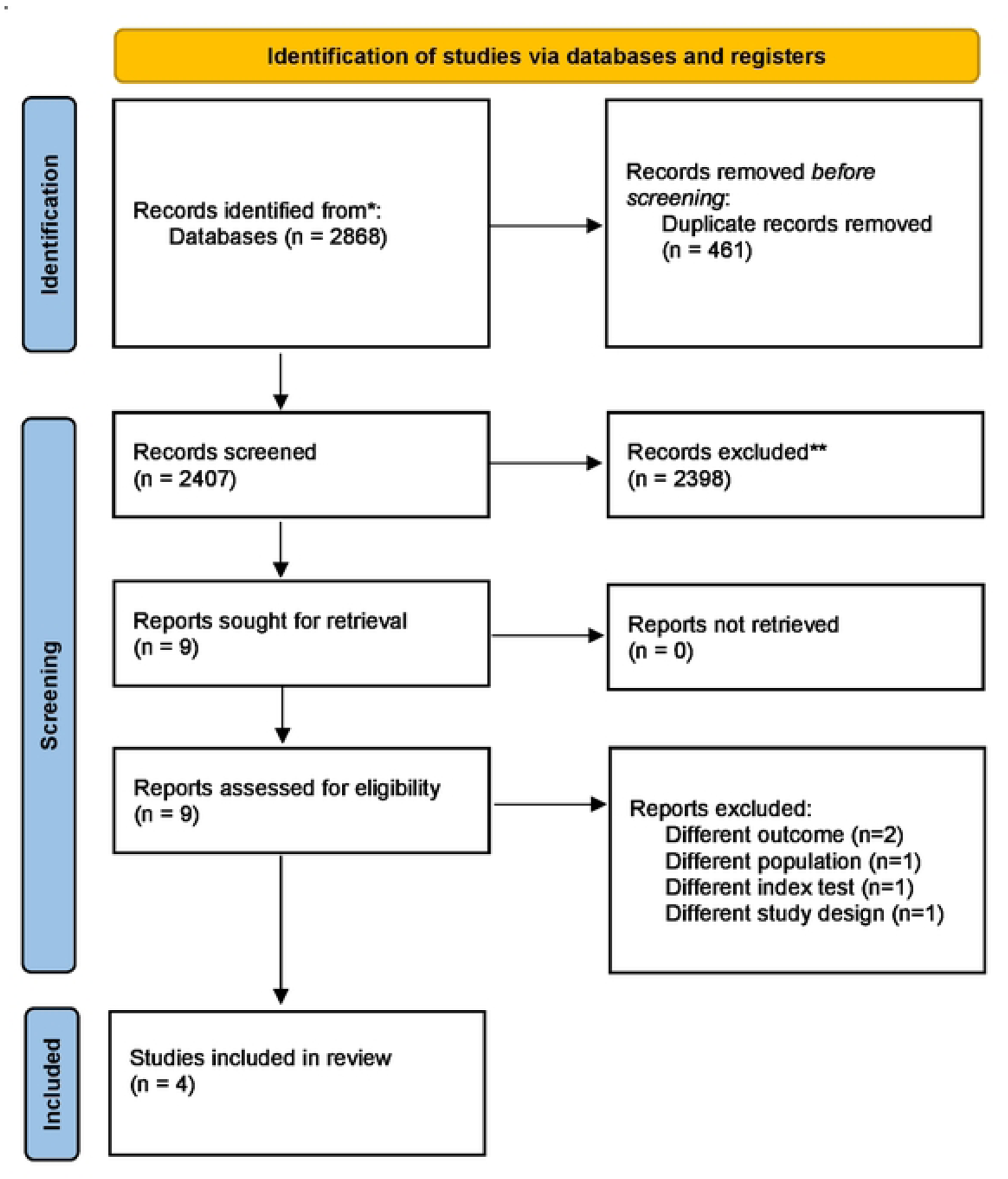
PRISMA flow diagram indicating the process of inclusion and exclusion of studies.

### Included and excluded studies

We excluded five studies (**Table S2**) because of the different populations, reference tests, or outcomes (i.e., neither provides DTA measures nor data to calculate them; **Table S2**). **Table 1** summarizes the characteristics of the four included studies (22–25). These studies were conducted in the United States (U.S.) (22,25), United Kingdom (U.K.) (22), Brazil (23), Bangladesh (24), and Thailand (25). In total, they included 3,122 (30% thresholds of the StandG6PD-BS), 2,371 (70%), and 2,371 (80%) participants for each test analysis, respectively. All studies were cross-sectional DTA, two included healthy adults (≥ 18 years old) (22,25), one included participants older than two years (23), and three studies included individuals with known G6PD status (23–25). The reference test used in all the studies was spectrophotometry for the G6PD kit (Pointe Scientific).

**Table 1.** Characteristics of included studies.

| First Author | Year | Study type | Countries | Study context | Inclusion criteria | Sample type | Threshold | N for test operative characteristics (2x2 tables) |  |  |  |  | Reference (index) test | Gold Standard | Observation | Funding |
| --- | --- | --- | --- | --- | --- | --- | --- | --- | --- | --- | --- | --- | --- | --- | --- | --- |
|  |  |  |  |  |  |  |  | TP | FN | FP | TN | Total |  |  |  |  |
| Pal (21) | 2021 | Cross-sectional DTA | U.S. (three clinical centers) U.K. | Healthy volunteers blood donors in different clinical centers | Healthy adults, ≥ 18 years old. They signed written informed consent. | Venous | 30% | 56 | 0 | 26 | 708 | 790 | STANDARD™ G6PD (SD Biosensor) | Spectrophotometry for G6PD using the kit Pointe Scientific | None | UK's FCDO, Bill & Melinda Gates fdn. |
|  |  |  |  |  |  |  | 70% | 82 | 1 | 49 | 658 | 790 |  |  |  |  |
|  |  |  |  |  |  |  | 80% | 85 | 16 | 58 | 631 | 790 |  |  |  |  |
| Zobrist (22) | 2021 | Cross-sectional DTA | Brazil | Febrile patients seeking care at the Manaus (924) and Porto Velho clinics (812) | Participants ≥ 2 years old plus 69 belonged through an enriched sample for G6PD known status. | Venous | 30% | 56 | 0 | 23 | 1583 | 1662 | STANDARD™ G6PD (SD Biosensor) | Spectrophotometry for G6PD using the kit Pointe Scientific | Measurement of temperature and humidity data at the time of testing that can affect PoC enzymatic activity test | UK's FCDO, Bill & Melinda Gates fdn., the National Institutes of Health |
|  |  |  |  |  |  |  | 70% | 31 | 1 | 31 | 848 | 911 |  |  |  |  |
|  |  |  |  |  |  |  | 80% | 41 | 20 | 21 | 829 | 911 |  |  |  |  |
| Alam (23) | 2018 | Cross-sectional DTA | Bangladesh | Participants were recruited in the Chittagong Hill Tracts districts (south-eastern) | Convenience sample from a cohort of adult with known G6PD status. | Venous | 30% | 30 | 0 | 0 | 76 | 106 | STANDARD™ G6PD (SD Biosensor) | Spectrophotometry for G6PD using the kit Pointe Scientific | Review of test results changes in blood samples stored at room temperature (24 to 26°C) and 4°C. | Wellcome Trust, Australian DFAT, Bill & Melinda Gates fdn. |
|  |  |  |  |  |  |  | 70% | 48 | 3 | 8 | 47 | 106 |  |  |  |  |
|  |  |  |  |  |  |  | 80% | 58 | 3 | 9 | 36 | 106 |  |  |  |  |
| Pal (24) | 2018 | Cross-sectional DTA | U.S. | Healthy volunteers blood donors in two clinical centers (New York and Miami) | Healthy adults, ≥ 18 years old, with an African American origin. | Fresh venous blood | 30% | 84 | 0 | 10 | 320 | 414 | STANDARD™ G6PD (SD Biosensor) | Spectrophotometry for G6PD using the kit Pointe Scientific (for Thailand, the Trinity Biotech assay was used). | None | UK's DFID, Bill & Melinda Gates fdn., MORU-Wellcome Trust |
|  |  |  |  |  |  |  | 70% | 105 | 5 | 9 | 295 | 414 |  |  |  |  |
|  |  |  |  |  |  |  | 80% | 115 | 6 | 40 | 253 | 414 |  |  |  |  |
|  |  | Cross-sectional DTA | Thailand | Participants attended two clinical centers located north and south of Mae Sot that serve a migrant population of Burman and Karen ethnic groups. | Adults with known G6PD status joined the study to achieve a convenience sample. | Frozen venous blood | 30% | 54 | 0 | 5 | 91 | 150 |  |  |  |  |
|  |  |  |  |  |  |  | 70% | 96 | 5 | 9 | 40 | 150 |  |  |  |  |
|  |  |  |  |  |  |  | 80% | 103 | 4 | 11 | 32 | 150 |  |  |  |  |

Abbreviations: TP: True Positives; FN: False Negatives; FP: False Positives; TN: True Negatives; G6PD: Glucose-6-Phosphate Dehydrogenase; DTA: Diagnostic Test Accuracy; U.S.: United States of America; U.K.: United Kingdom; FCDO: Foreign, Commonwealth & Development Office; fdn.: foundation; DFAT Department of Foreign Affairs and Trade, DFID: Department for International Development; MORU: Mahidol Oxford Tropical Medicine Research Unit

### Risk of Bias (RoB)

Two studies were judged as high (24,25), and two as low RoB in domain one (22,23) (patient selection) of the QUADAS-2 tool. One study was judged high (24), two unclear (22,25), and one low RoB in domain four (23) (flow and timing). All studies had low RoB and applicability concerns in domains two and three (reference test and standard). This information is presented as a table and diagram using the QUADAS-2 tool resources (**Figure 2**).

**Figure 2.**
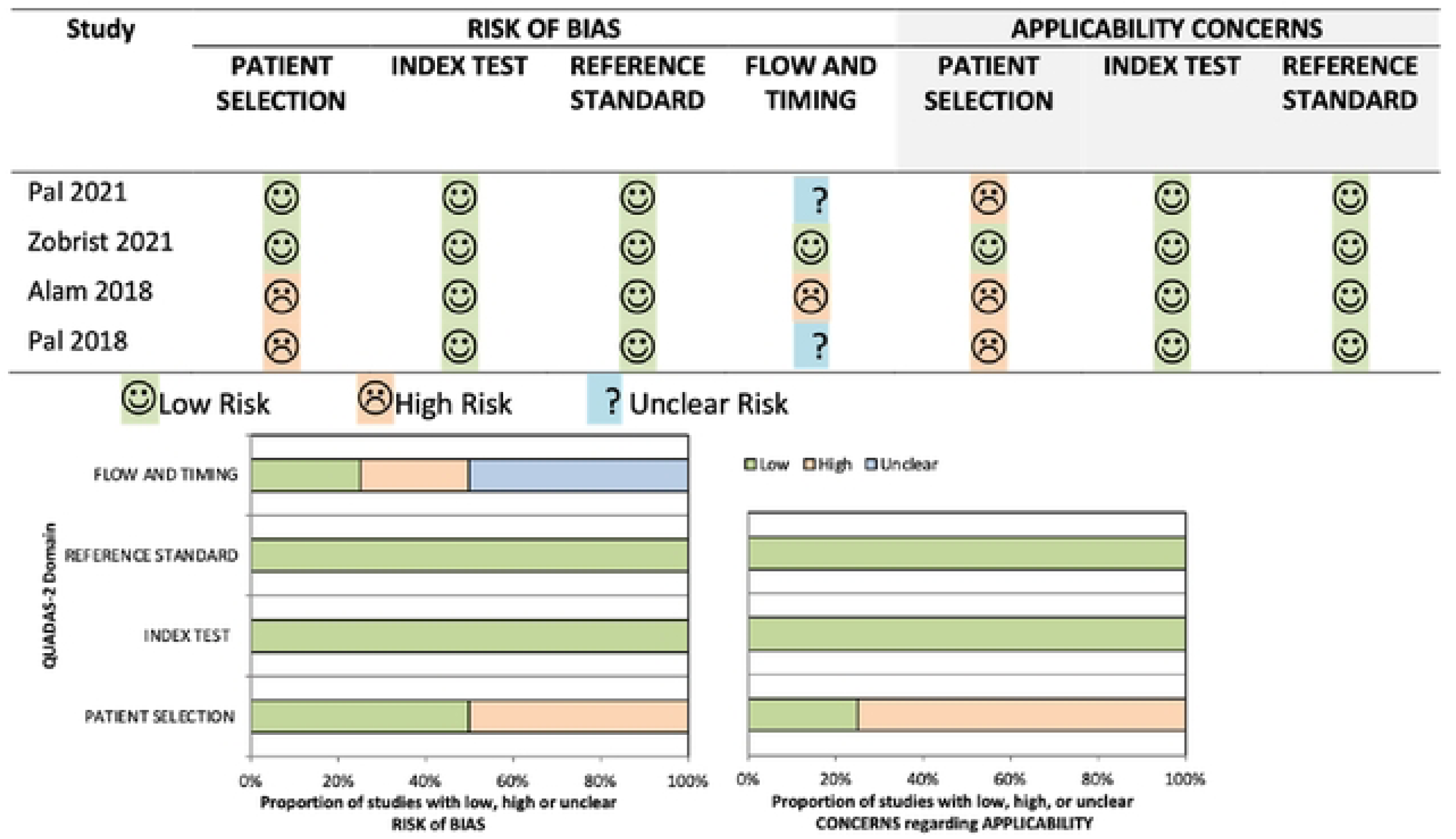
RoB summary judgements about each included study using QUADAS-2 tool.

### Diagnostic test accuracy from primary studies and pooled data

Sensitivities and specificities from primary studies are presented in **Figure 3**. Sensitivity ranged from 91% to 99%, whereas specificities were from 89% to 97% for 30% and 80% thresholds, respectively. **Figure 3** displays the crosshair plot (sensitivity vs. false positive rates) for the three thresholds. Positive and negative LR from studies are presented in **Figure S1**. Pooled DTA measures for each G6PD activity threshold are described (**Table 2**). Pooled results showed high DTA measures for all the thresholds being better for lower than the highest threshold. Positive and negative LR ranged from 8.2 to 35.3 and 0.009 to 0.106. The CoE for sensitivity and specificity was high for 30% and 70% thresholds, but for 80% threshold sensitivity was low and specificity was moderate due to concerns in the RoB, indirectness, inconsistency, and imprecision (See **Table S3**). We found low, moderate, and high heterogeneity in the results for the 30%, 70%, and 80% thresholds, respectively (**Figure 4 and 5)**.

**Figure 3.**
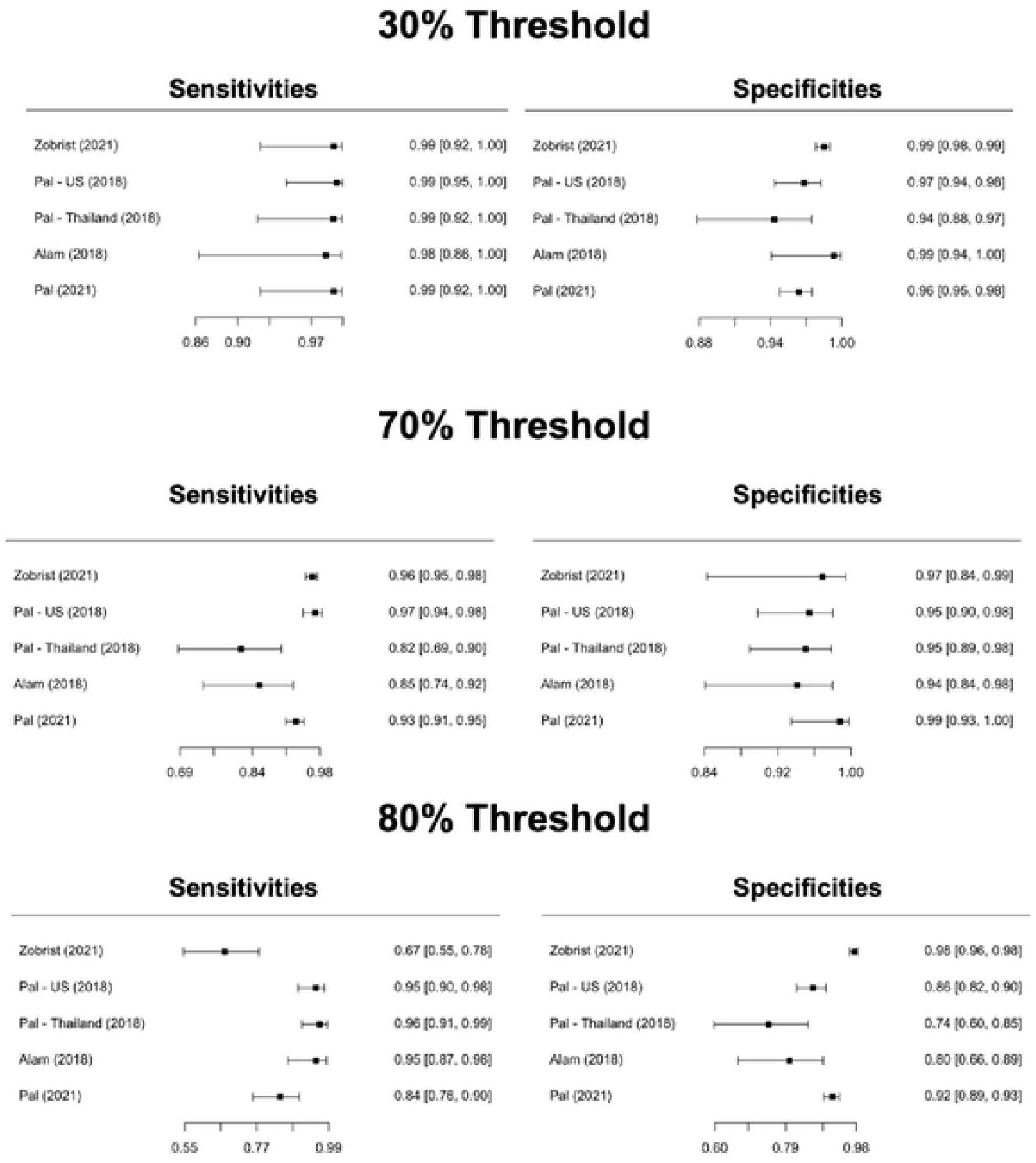
Pooled Sensitivity and Specificity for Standard^TM^ G6PD (SD Biosensor) in venous blood samples.

**Figure 4.**
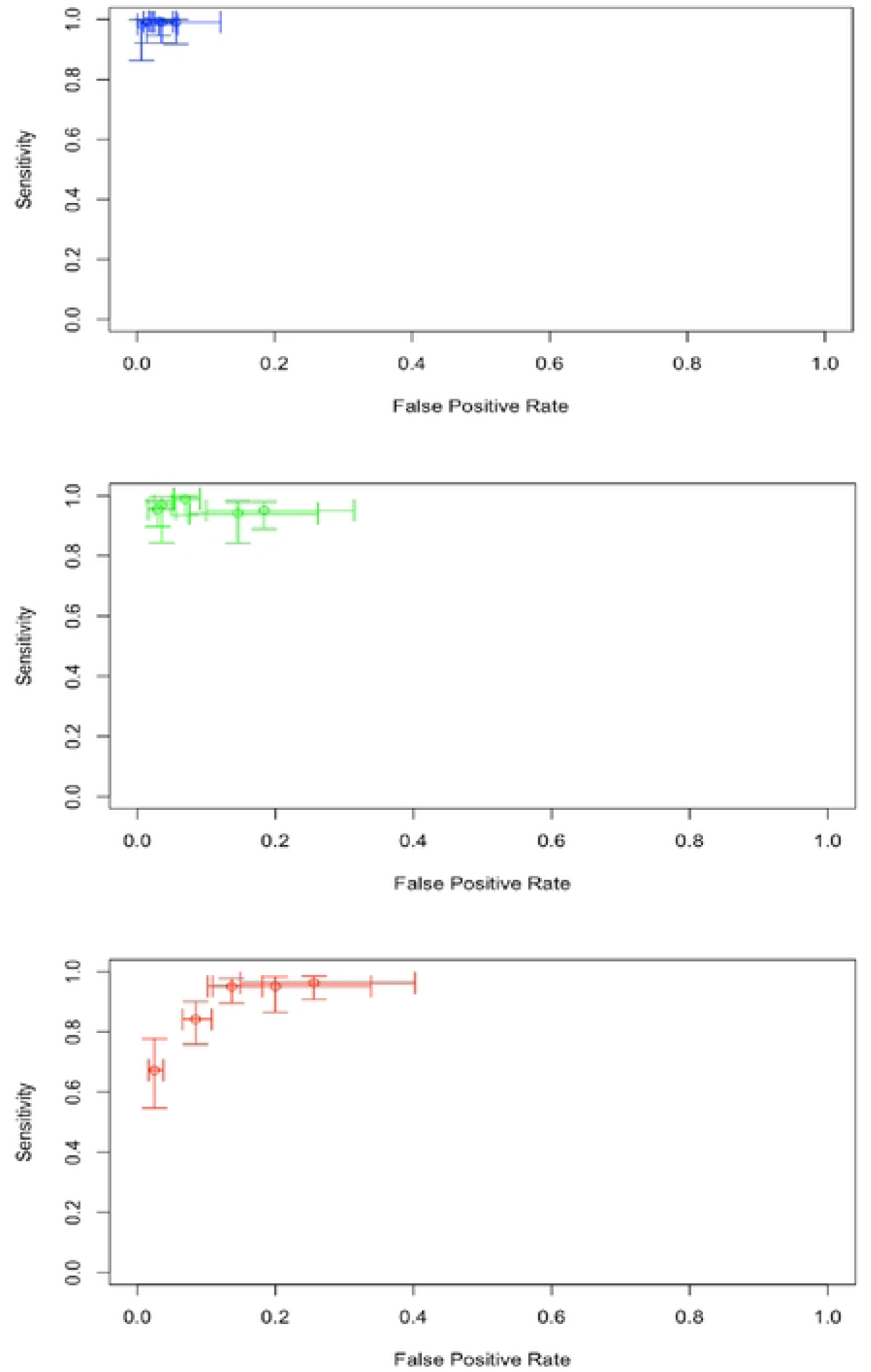
Crosshair plot for each threshold defined by the Standard^TM^ G6PD (SD Biosensor) test.

**Figure 5.**
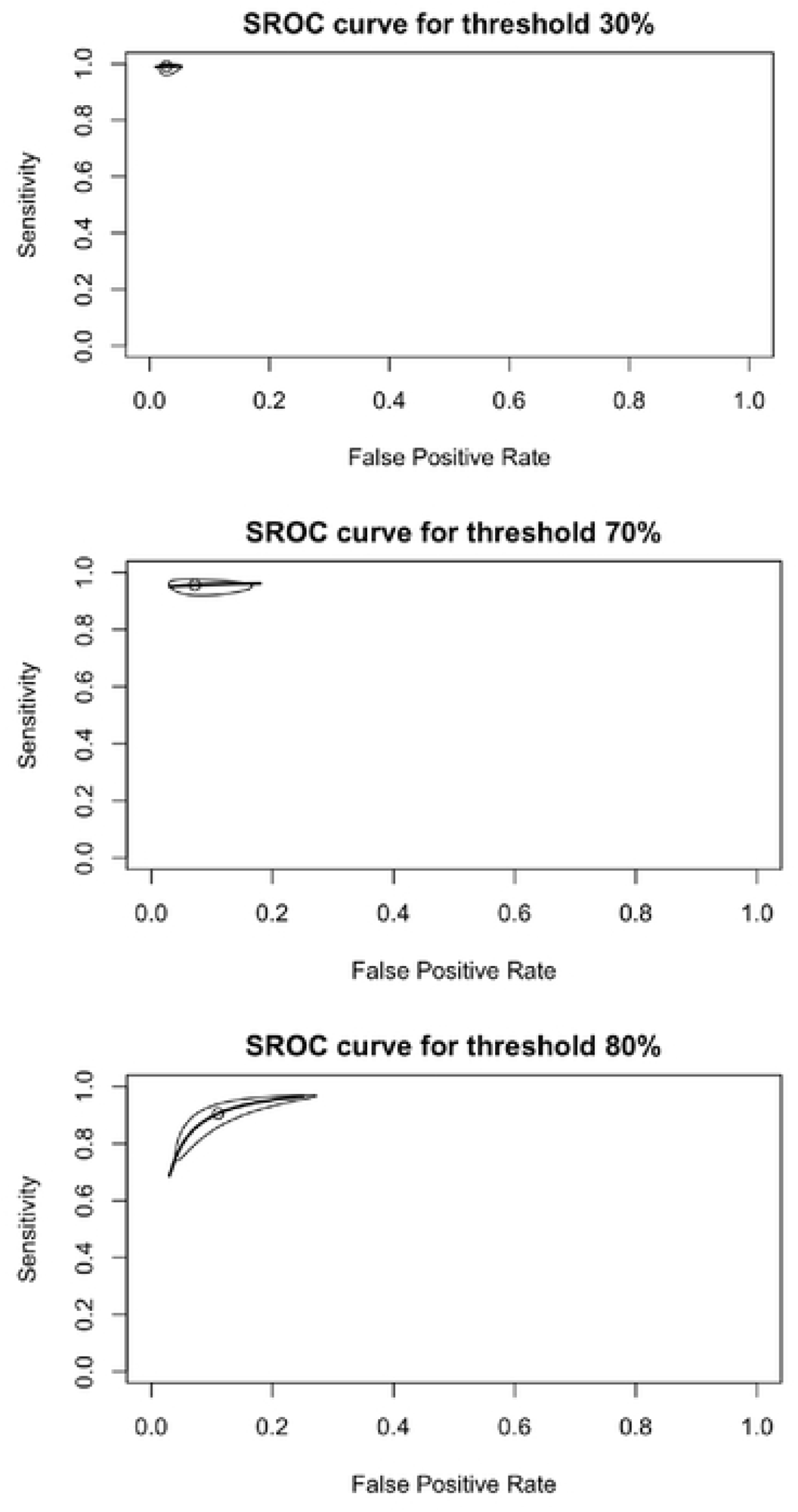
SROC curve (bivariate model) for different Standard G6PD (SD Biosensor) thresholds in venous blood samples.

**Table 2.** Pooled DTA values for each threshold defined by Standard^TM^ G6PD (SD Biosensor) test.

| **G6PD activity threshold** | **Sensitivity (95%CI) & CoE** | **Specificity (95%CI) & CoE** | **+LR (95%CI)** | **-LR (95%CI)** |
| --- | --- | --- | --- | --- |
| 30% | 99.1% (96.9%-99.7%)<br>(4 studies, 3122 participants) | 97.4% (95.2-98.4%)<br>(4 studies, 3122 participants) | 35.3<br>(20.7-60.5) | 0.009<br>(0.003-0.031) |
|  | High ⊕⊕⊕⊕ | High ⊕⊕⊕⊕ |  |  |
| 70% | 95.7% (92.9-97.4%)<br>(4 studies, 2371 participants) | 92.8% (85.8-96.5%)<br>(4 studies, 2371 participants) | 13.2<br>(6.8-26.5) | 0.046<br>(0.030-0.073) |
|  | High ⊕⊕⊕⊕ | High ⊕⊕⊕⊕ |  |  |
| 80% | 90.5% (78.4-96.2%)<br>(4 studies, 2371 participants) | 89.0% (76.9-95.1%)<br>(4 studies, 2371 participants) | 8.2<br>(4.1-16.0) | 0.106<br>(0.049-0.227) |
|  | Low ⊕⊕○○ | Moderate ⊕⊕⊕○ |  |  |

*Abbreviations: CI: confidence interval, LR: likelihood ratio, +: positive, -: negative, CoE: Certainty of the Evidence*

## Discussion

In this systematic review, we included four studies evaluating 7,864 patients and found that StandG6PD-BS is a semi-quantitative PoC test with high sensitivity and specificity values to detect G6PDd at the different G6PD activity thresholds recommended by the manufacturer (30%, 70%, and 80%). The test showed better performance and CoE for 30% and 70%, compared with the 80% threshold. Likewise, the heterogeneity in the results was low, moderate, and high at 30%, 70%, and 80% thresholds, respectively.

The included studies were conducted in both high and low-middle-income countries, and participants were predominantly healthy adults with or without known G6PD status. This allows the decision-makers to assess the potential impact of implementing the StandG6PD-BS PoC test in the general population, particularly for uncomplicated malaria by *P. vivax*. The evidence suggests that introducing this test in tropical and subtropical malaria-endemic countries with *P. vivax* high transmission regions would be both feasible and desirable and could provide numerous benefits (5,7,26). For instance, a recent study in Brazil (26) found that combining TQ treatment with the StandG6PD-BS test improved the response to radical cure treatment by enhancing adherence, reducing relapses, and increasing protection against new *P. vivax* infections.

The literature describes factors related to lower test accuracy that may not be explained just by the PoC test performance. A recent controlled study suggested that the StandG6PD-BS test performed well in venous blood, exhibiting good repeatability and inter-laboratory reproducibility (27). However, the same study found that the reliability of the test was poor in discriminating between intermediate and low G6PD activities in lyophilized samples (27), emphasizing the need for further research in field-based scenarios. This study found similar estimates for sensitivity and specificity as reported by Addisu et al. (15). However, our work is the first to conduct a complete systematic review approach through a comprehensive search and selection, a random model meta-analysis, risk of bias assessment, and certainty of evidence using the GRADE approach and following the methods recommended by the Cochrane collaboration.

In our approach, we pooled only results of venous blood samples because other sources, such as capillary and lyophilized blood, provided limited results due to small sample sizes in the studies and differences in the specimen collection and lyophilization methods. Nevertheless, some evidence suggests that capillary and lyophilized blood samples could drive less accuracy in the PoC test results (22,27–29).

In the included studies, only one (25) used frozen venous blood to run the StandG6PD-BS test showing slightly less DTA than the fresh venous blood samples (25). This could be due to the small number of participants, the specimen collection, or the storage method. An alternative cause is the study population (Thailand), which carries the G6PDd Mahidol phenotype with moderate enzymatic activity (30-70%). However, the StandG6PD-BS sensibilities and specificities at those levels are still around 90% (22,27,30).

The current evidence strongly supports the implementation of the PoC test for G6PD activity. At the individual level, it will enable the safe treatment of more patients with deficient and intermediate G6PD activity, diminishing the risk of recurrent malaria and acute hemolytic anemia (5,7,31,32). At the healthcare systems level, it could reduce the associated costs and the burden on transfusion services by reducing the number of hemolytic crises caused by PQ or TQ in individuals with unknown G6PD status and malaria treatment (5,7,31,33). Additionally, this could impact parasite transmission rates when combined with other interventions (5,7,31). Given that up to 50% of the patients with *P. vivax* malaria may experience relapses (5,7), administering radical cure with PQ or TQ is essential for stopping morbidity-related and community transmission (5,7,26,32).

Our work has several strengths. This systematic review with meta-analysis is the first to evaluate the DTA of the StandG6PD-BS test. Moreover, we followed state-of-the-art methodologies for conducting DTA studies, as recommended by the Cochrane Handbook for Systematic Reviews of Diagnostic Test Accuracy (34). We prepared the final report of this review following the recommendations by the PRISMA-DTA statement (35). We also provided information on the pooled sensitivity and specificity for each threshold, which can facilitate decision-making in different clinical scenarios.

We also recognize some limitations. First, the number of studies was low, preventing us from performing additional analyses, such as sensibility analysis. Additionally, the quality of some studies was not optimal, although our analysis is robust enough to show adequate pooled DTA measures for venous blood samples. Future studies using capillary and lyophilized blood, along with field-based studies, are needed to determine the appropriate usage of this test. A study on barriers and facilitators for G6PD test implementation (36) identified three main barriers: perceived low risk of hemolysis, wrong perception of *P. vivax* malaria as a benign condition, and the cost of routine testing as part of the healthcare attention of malaria patients. A study conducted in Brazil on the operational challenges associated with pragmatic G6PD testing (37) found that the StandG6PD-BS PoC test was well accepted by both healthcare professionals and patients and can be performed at malaria treatment units in the Brazilian Amazon to inform treatment decisions with PQ. However, the authors found limitations linked to technical and cultural aspects that should be addressed when expanding screening to larger areas (37).

## Conclusion

StandG6PD-BS is a PoC test with high sensitivity and specificity to detect G6PDd at the different thresholds recommended by the manufacturer (30%, 70%, and 80%). Implementing this kind of test in malaria-endemic areas can lead to early diagnosis of G6PDd, help to prevent hemolytic episodes triggered by PQ or TQ, and potentially impact malaria transmission.

## Data Availability

Data for the analyses was obtained from published studies. Additional data are available on request from the authors.

## Acknowledgments

We acknowledge Dr Lina Gaviria (Haematologist, Universidad de Antioquia, Colombia), for their contribution to the conceptualization of this study; Marina Figueira, and Camilo Manchola (Global Health Strategies), Jonathan Novoa (Medicines for Malaria Venture), and Jamil Barton and Reina Jara (Program for Appropriate Technology in Health) for their contribution in this study.

## Data Availability Statement

The data are available on request from the authors.

## Author’s contributions

**Conceptualization:** JCM, IDF.

**Data curation**: JCM, VVM.

**Formal analysis**: JCM, VVM, IDF, MLP

**Funding acquisition:** IDF, DFP.

**Investigation:** IDF, DFP, JCM, VVM.

**Methodology:** JCM, VVM.

**Project administration:** JCM.

**Resources:** IDF.

**Software:** JCM

**Supervision:** IDF, DFP.

**Visualization:** JCM, VVM.

**Writing – original draft:** VVM, JCM.

**Writing – review & editing:** JCM, VVM, DFP, IDF, MLP.

