## Supplementary material for "Diagnostic accuracy of the Point-of-Care Standard G6PD test™ (SD Biosensor) for Glucose-6-phosphate dehydrogenase deficiency: a systematic review of the literature": Table S

**Supplementary files**

**Table S1. Search strategy**

**PubMed-MEDLINE**

"glucosephosphate dehydrogenase deficiency"[MeSH Terms] OR "favism"[MeSH Terms] OR "glucose-phosphate dehydrogenase deficienc*"[Title/Abstract] OR "glucosephosphate dehydrogenase deficienc*"[Title/Abstract] OR "glucosephosphate dehydrogenase activity"[Title/Abstract] OR "glucose-phosphate dehydrogenase activity"[Title/Abstract]"glucose-6-phosphate dehydrogenase deficienc*"[title/abstract]or "glucose-6-phosphate dehydrogenase activit*"[Title/Abstract] OR "favism"[Title/Abstract] OR "G6PD deficienc*"[Title/Abstract] OR "G6PD activity*"[Title/Abstract] OR "G6PD-deficienc*"[Title/Abstract] OR "Enzymopath*"[Title/Abstract]) AND ("diagnos*"[MeSH Terms] OR "diagnos*"[tiab] OR "sensitivity and specificity"[MeSH Terms] OR "sensitiv*"[Title/Abstract] OR "specificit*"[Title/Abstract] OR "Point-of-Care Testing”[MeSH Terms] OR “Point-of-Care Test*"[Title/Abstract])

**OVID EMBASE**

1.	exp Glucosephosphate Dehydrogenase Deficiency/

2.	exp Favism/

3.	glucose-phosphate dehydrogenase deficienc$.mp.

4.	glucosephosphate dehydrogenase deficienc$.mp.

5.	(glucose-phosphate dehydrogenase activity or glucosephosphate dehydrogenase activity).tw.

6.	glucose-6-phosphate dehydrogenase deficienc$.mp.

7.	glucose-6-phosphate dehydrogenase activit$.mp.

8.	favism.mp.

9.	g6pd.tw.

10.	Enzymopath$.mp.

11.	1 or 2 or 3 or 4 or 5 or 6 or 7 or 8 or 9 or 10

12.	sensitiv:.tw. or diagnostic accuracy.sh. or diagnostic.tw. or specificity.tw. or accuracy.tw. or (predictive and value$).tw.

13.	point-of-care test$.tw. or exp Point-of-Care Systems/

14.	12 or 13

15.	11 and 14

16.	exp animals/ not humans.sh.

17.	15 not 16

**Table S2.** Excluded studies.

| **First author** | **Year** | **Country** | **Study type** | **Reason for exclusion** | **Observations** |
| --- | --- | --- | --- | --- | --- |
| Ley (26) | 2022 | Indonesia | Repeated measures | Different outcome | This study aimed to determine the STANDARD^TM^ G6PD (SD biosensor) repeatability (assay precision when repeated under constant conditions) and reproducibility (assay precision under different conditions), not to assess DTA. |
| Gerth-Guyette (32) | 2021 | Brazil, Ethiopia  India | Quasi-experimental assay | Different population | This study seeks to assess how well end users can understand the STANDARD^TM^ G6PD (SD biosensor) workflow, result output, and label after training, not to assess DTA. |
| Brito-Sousa (33) | 2021 | Brazil | Mixed-methods study | Different index test | This study aimed to evaluate the routine implementation of the CareStart G6PD qualitative test at malaria treatment units (MTUs) of a municipality in the Brazilian Amazon (another index test). |
| Adissu  (34) | 2022 | Multiples countries | Retrospective analysis | Study design | Information not available for construction of 2x2 tables (even in primary studies). |
| Brito-Sousa  (31) | 2022 | Brazil | Operational Mixed-Methods Study | Different outcome | This study evaluated the operational challenges and acceptance for healthcare providers and patients of the STANDARD^TM^ G6PD (SD biosensor) testing. It did not have information about test performance. |

*Abbreviations: DTA: diagnostic test accuracy.*

**+Table S3.** GRADE Summary of findings (SoF) Tables

**Figure S1.** **Forest plots with Likelihood Ratio (LR)s from included studies for Standard^TM^ G6PD (SD Biosensor) in venous blood samples.** Forest Plot for displaying the positive LR and negative LR per study according to specific thresholds with their corresponding 95% CI*.*

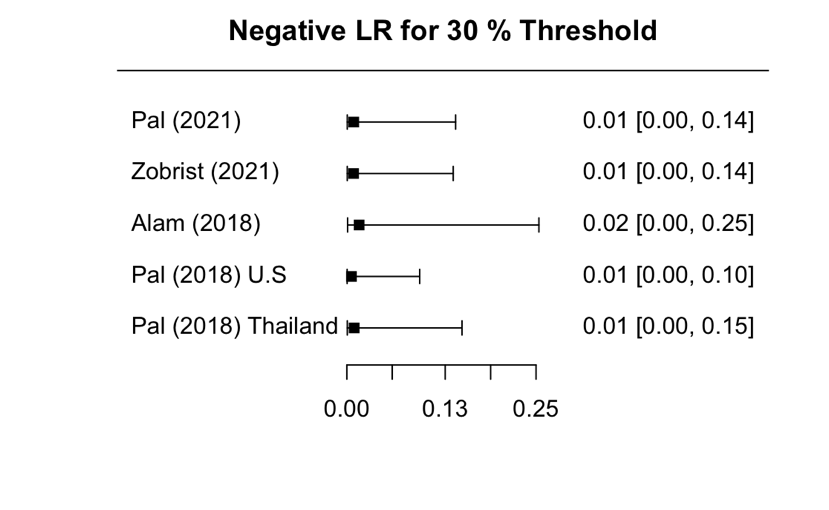

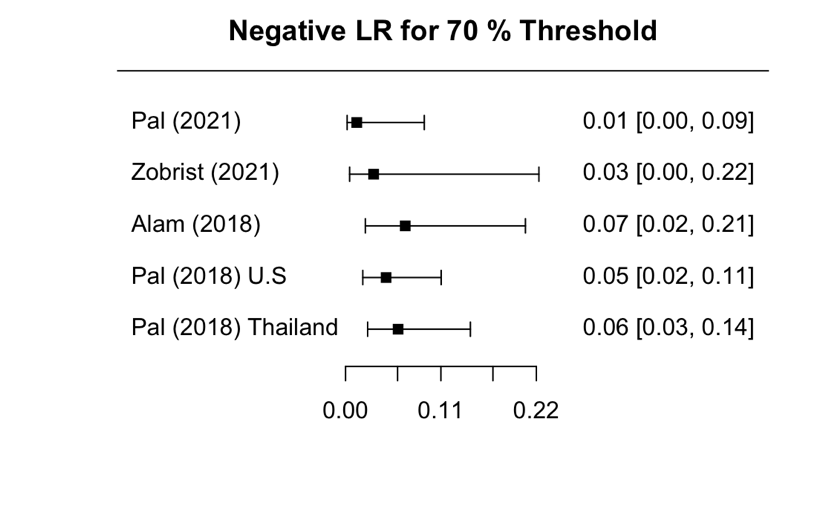

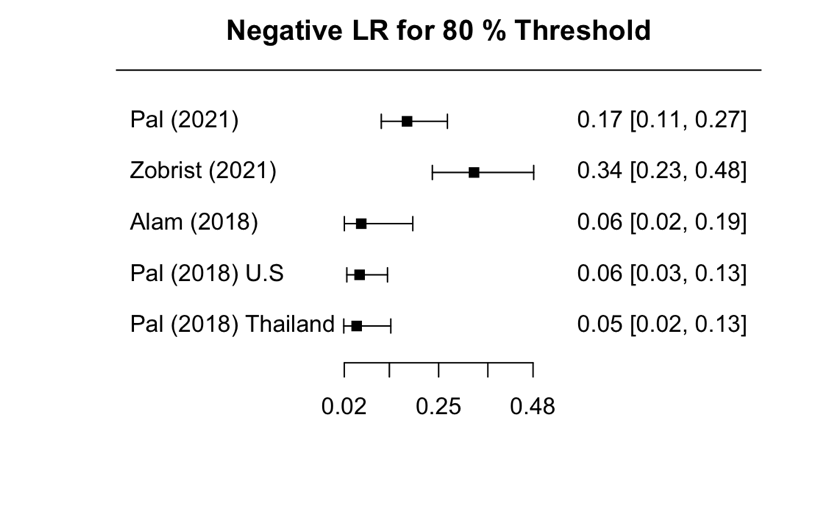

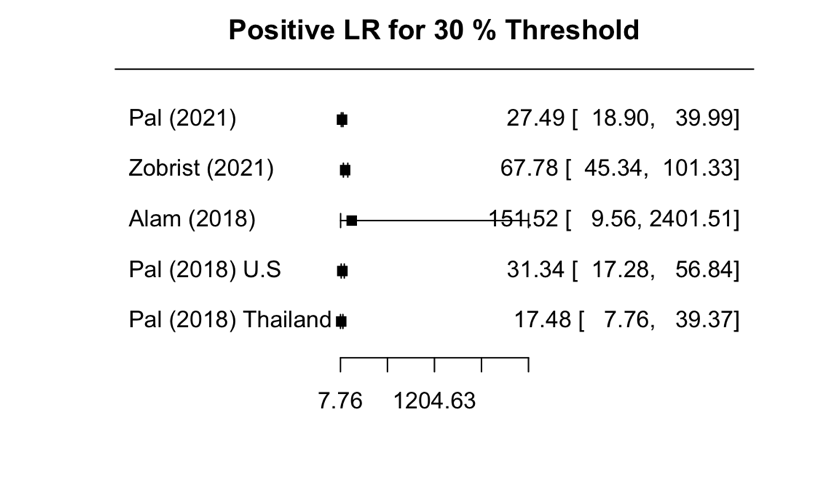

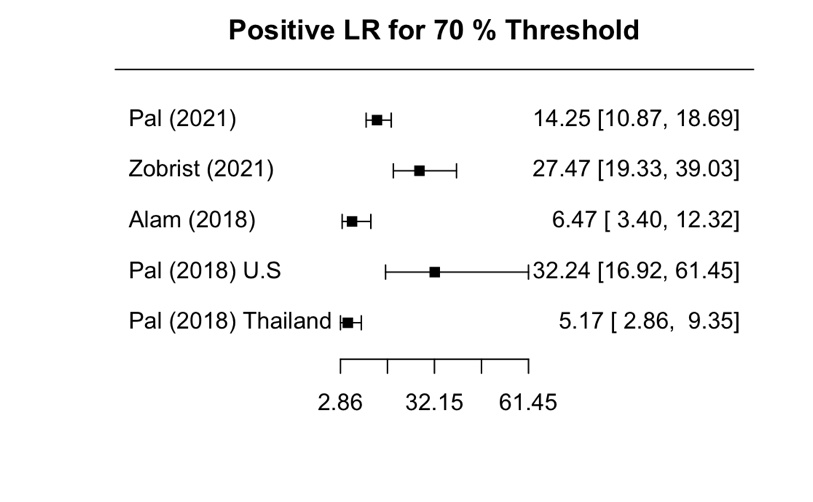

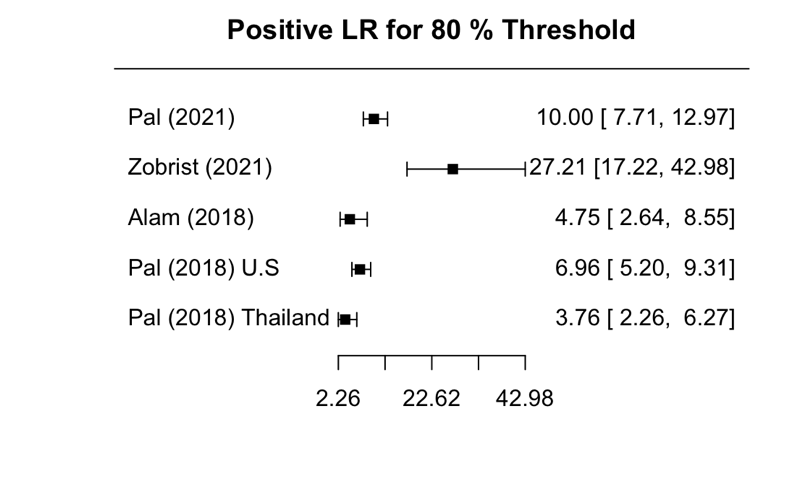
